# Impact of the COVID-19 Pandemic on Antimicrobial Resistance (AMR) Surveillance, Prevention and Control: A Global Survey

**DOI:** 10.1101/2021.03.24.21253807

**Authors:** Sara Tomczyk, Angelina Taylor, Allison Brown, Marlieke de Kraker, Tim Eckmanns, Aiman El-Saed, Majid Alshamrani, Rene Hendriksen, Megan Jacob, Sonja Löfmark, Olga Perovic, Nandini Shetty, Dawn Sievert, Rachel Smith, John Stelling, Siddhartha Thakur, Barbara Tornimbene, Ann Christin Vietor, Sergey Eremin, on behalf of the WHO AMR Surveillance and Quality Assessment Collaborating Centre Network

## Abstract

**Objectives:** The COVID-19 pandemic has had a substantial impact on health systems. The WHO Antimicrobial Resistance (AMR) Collaborating Centres Network conducted a survey to assess the effects of COVID-19 on AMR surveillance, prevention and control.

**Methods:** From October-December 2020, WHO Global Antimicrobial Resistance and Use Surveillance System (GLASS) national focal points completed a questionnaire including Likert-scales and open-ended questions. Data were descriptively analysed, income/regional differences were assessed, and free-text questions were thematically analysed.

**Results:** Seventy-three countries across income levels participated. During the COVID-19 pandemic, 67% reported limited ability to work with AMR partnerships; decreases in funding were frequently reported by low- and middle-income countries (LMICs; p<0.01). Reduced availability of nursing, medical and public health staff for AMR was reported by 71%, 69% and 64%, respectively, whereas 67% reported stable cleaning staff availability. The majority (58%) reported reduced reagents/consumables, particularly LMICs (p<0.01). Decreased numbers of cultures, elective procedures, chronically ill admissions and outpatients and increased intensive care unit admissions reported could bias AMR data. Reported overall infection prevention and control (IPC) improvement could decrease AMR rates, whereas increases in selected inappropriate IPC practices and antibiotic prescribing could increase rates. Most did not yet have complete data on changing AMR rates due to COVID-19.

**Conclusions:** This was the first survey to explore the global impact of COVID-19 on AMR among GLASS countries. Responses revealed universal patterns but also captured country variability. Although focus is understandably on COVID-19, gains in detecting and controlling AMR, a global health priority, cannot afford to be lost.

## Introduction

The coronavirus disease 2019 (COVID-19) pandemic has had a substantial impact on health systems globally, affecting the management of other health threats such as antimicrobial resistance (AMR). The World Health Organization (WHO) declared that AMR is one of the top ten global health threats and, although often more silent than the COVID-19 pandemic, it can have equally devastating consequences.^1^ From 2017-2019, the number of countries reporting AMR rates to WHO’s Global Antimicrobial Resistance and Use Surveillance System (GLASS) exponentially grew from 729 in 22 countries to more than 64,000 in 66 countries.^2^ However, the effects of the COVID-19 pandemic threatens the progress made and is thought to be having wide-reaching impacts on AMR surveillance, prevention and control efforts. Experts have highlighted the link between COVID-19 and AMR, indicating that certain changes such as increased antibiotic use could drive an increase in AMR; while other activities such as improved infection prevention and control (IPC) might reduce AMR rates.^3-6^ This underlines the importance of maintaining AMR surveillance to monitor trends during the COVID-19 pandemic.

The WHO AMR Surveillance and Quality Assessment Collaborating Centres Network is a global network of institutes with expertise in AMR, health care-associated infections, and antimicrobial consumption. It aims to support WHO’s efforts to combat AMR through the development and implementation of AMR surveillance and related activities.^7^ We conducted a survey among countries enrolled in GLASS to assess the global effects of COVID-19 on AMR surveillance, prevention and control, focusing on challenges as well as opportunities.

## Materials and methods

A structured questionnaire was developed with expert input from Network members (Supplementary data). The WHO health system building blocks framework was considered to ensure that the impacts of COVID-19 on different health system areas were comprehensively addressed.^8^ Accordingly, the questionnaire consisted of compulsory Likert-scale questions to assess the impacts of COVID-19 in ten topic areas (i.e. 2-10 questions per topic area): Funding for AMR activities; Partnerships and oversight for AMR activities; Diagnostics and laboratory testing for AMR; Laboratory supplies and equipment for AMR activities; Availability of staff for AMR activities; AMR data information systems; Patient-case mix; IPC practices; Antibiotic consumption; and AMR rates (Supplementary data). Likert-scale responses included “Large decrease”, “Moderate decrease”, “No impact”, “Moderate increase”, “Large increase”, and “Do not know.” To further explore country experiences, each topic area ended with an optional open-ended question and three optional open-ended questions were included at the end of the questionnaire. Upon approval from the data protection office of the Network coordinator (Robert Koch Institute, Berlin, Germany), the questionnaire was programmed using the online Voxco survey software including validity and completeness checks.

From October-December 2020, the questionnaire link was sent to the national focal points of all countries enrolled in GLASS. Each GLASS national focal point was asked to consider input from their relevant country experts (e.g. epidemiology, clinical, laboratory, IPC) and submit one compiled response per country. There was active follow-up with reminders, and countries which submitted more than one response were requested to indicate their final response.

The data collected were descriptively analysed using the statistical programme R (version 4.0.3). Completed Likert-scale responses were graphically displayed for each of the ten topic areas. Differences in responses between countries according to WHO regions and World Bank income levels^9^ were assessed using the Fisher’s exact test and significant differences (p<0.05) were reported. Free-text questions were reviewed to identify specific themes, coded accordingly and considered in relation to the corresponding topic area Likert-scale findings (Supplementary data). If minor typographical errors were corrected in quotations for comprehension, this was indicated with “sic”.

## Results

A total of 73 countries responded to the survey, corresponding to 75% of countries enrolled in GLASS at the time of the survey (Table 1). The regional and income distribution of survey respondents was similar to those in GLASS, including 16% (12/73) low-income, 23% (17/73) lower middle-income, 21% (15/73) upper middle-income and 37% (27/73) high-income countries (Table 1). The median number of countries providing a response for each mandatory question was 66 (i.e. incompleteness included selection of “Do not know”).

**Table 1.** Descriptive characteristics of survey respondents (“Survey”; N=73) compared to total countries participating in GLASS at the time of the survey (“GLASS”; N=98)

| Characteristic | Survey |  | GLASS |  |
| --- | --- | --- | --- | --- |
|  | N | % | N | % |
| **Income level** |  |  |  |  |
| Low-income | 12 | 16.4% | 17 | 17.3% |
| Lower Middle-income | 17 | 23.3% | 27 | 27.6% |
| Upper Middle-income | 15 | 20.5% | 20 | 20.4% |
| High-income | 27 | 37.0% | 34 | 34.7% |
| Unknown* | 2 | 2.7% |  |  |
| Total | 73 | - | 98 | - |
| **Region** |  |  |  |  |
| African Region (AFR) | 16 | 21.9% | 26 | 26.5% |
| Region of the Americas (AMR) | 5 | 6.8% | 6 | 6.1% |
| South-East Asia Region (SEAR) | 7 | 9.6% | 11 | 11.2% |
| European Region (EUR) | 20 | 27.4% | 25 | 25.5% |
| Eastern Mediterranean Region (EMR) | 18 | 24.7% | 21 | 21.4% |
| Western Pacific Region (WPR) | 5 | 6.8% | 9 | 9.2% |
| Unknown* | 2 | 2.7% |  |  |
| Total | 73 | - | 98 | - |

\*Respondent chose not to provide their country name

### Funding for AMR activities

Among those that responded, less than half of countries reported decreases in funding for AMR activities at the national (29/70; 41%) or local/facility level (30/69; 43%) (Figure 1a). Decreases in national and local funding were reported more frequently by low-(9/10; 9/10) and middle-income (15/30; 16/30) countries compared to high-income (3/27; 3/26) countries, respectively (p<0.01). Decreases were also more frequently reported by the African and Eastern Mediterranean regions compared to other regions (p<0.01). In the free-text questions (Table 2), various countries reported that funding was prioritized for COVID-19 over AMR. This ranged from selected low-income countries who reported being dependent on external funding for AMR that was impacted by COVID-19 to high-income countries that reported more indirect effects which reduced resources for AMR activities. In contrast, one middle-income country reported that the COVID-19 pandemic allowed them to secure additional AMR funding, and another was able to purchase resources for overall IPC with COVID-19 funds.

**Table 2.** Selected free-text reports by countries according to topic area and income level.

| Topic | Income level | Selected illustrative quotations |
| --- | --- | --- |
| **Funding for AMR activities** | Low | "Previously, there was a small fund from the WHO country office, but during the COVID-19 pandemic, all funding and activities for AMR stopped till now...During the last 10 months, all focus is on COVID-19 and there is no support for AMR by governments and non-governmental organizations." |
|  | Lower-middle | "AMR surveillance activities for national and local levels moderately decreased due to the large amount of funding allocated to laboratory services and treatment for covid-19 patients." |
|  | Upper-middle | "There was no funding for AMR surveillance at the national level before the pandemic." |
|  | High | "Indirectly, we may conclude that there is a decrease in AMR support as most microbiologists and epidemiologists were mobilized for COVID diagnostics." |
| **Partnerships and oversight for AMR activities** | Low | "Due to the mobility restrictions, activities focused on AMR stopped. Sometimes, we try to use distanced calls but no success. Internet connection is very limited in the country, it was difficult to reach each site [sic]." |
|  | Lower-middle | "The WHO country office had requested a consultant to support our efforts to develop the AMR master plan, but due to the COVID-19 outbreak, all those plans failed [sic]."<br>"COVID-19 has created platforms for new partnerships and collaborations because of the link in Infection Prevention interventions, e.g. Water and Sanitation and Hygiene (WASH)." |
|  | Upper-middle | "The COVID-19 pandemic crisis and the issuance of some strict measures to confront the Corona epidemic, including the imposition of a complete curfew, led to poor communication with partners." |
|  | High | "More people and organisations have found each other, more possibilities regarding data exchange." |
| **Diagnostics and laboratory testing for AMR** | Low | "The schedules which had been made to train staff were stalled by the COVID-19 Pandemic. Laboratory turnaround time rose due to less staff levels than usual on the microbiology benches [sic]." |
|  | Lower-middle | "The laboratory network was strengthened in [our country] as part of the COVID-19 response and this will positively impact the AMR surveillance network. The decision makers are now very sensitized to labs issues [sic]." |
|  | Upper-middle | "Patients avoided visiting hospitals as they were afraid to be in close contact to the healthcare personnel and inpatients. This resulted in a decrease of patient visits and microbiological orders [sic]." |
|  | High | "Diagnostic pathology activity in microbiology laboratories...has declined when compared with the steep rise in testing work associated with detection of SARS-COV-2."<br>"Whole genome sequencing (WGS) activity on antibiotic resistance strains has decreased because of the availability of WGS machines (reserved for Covid), of molecular reagents and of staff (half team and staff rotation) [sic]." |
| **Laboratory supplies and equipment for AMR activities** | Low | "Because there has been a drop in samples being analysed and patients seen facilities, this has caused a reduction in the amount of resources spent on supplies and consumables [sic]." |
|  | Lower-middle | "There was no impact of COVID-19 on laboratory supplies and equipment for AMR activities as there was no functional surveillance during the COVID-19 pandemic." |
|  | Upper-middle | "During Covid-19 lock out, we had many difficulties to import reagents, equipment and some parts in order to repair the equipment [sic]." |
|  | High | "There was some impact on nucleic acid amplification-related work rather than standard culture and antimicrobial susceptibility testing. Assays detecting resistance genes via nucleic acid amplification were in some cases delayed due to the availability of PCR platforms which were in use mostly for SARS-COV-2 RNA detection." |
| **Availability of staff responsible for AMR activities** | Low | "Human resources has been one of the areas affected due to covid-19 responses, a lot of staff have been pulled to support covid-19 and this leads to no activities and actions done." |
|  | Lower-middle | "In general, most of health staff (doctors, nurses, lab staff, etc.) were called to respond activities of Covid-19 emergency, affecting the availability of these professionals for AMR activities [sic]." |
|  | Upper-middle | "We had a 2 [moderate impact] in the availability of health professionals in several places and a great increase in the need for professionals during the beginning of the pandemic. Thus, the government supported the hiring of professionals through a specific program that identified non-employed professionals, created a national register of professionals and local demand, and then allocated these professionals where they were most needed." |
|  | High | "Public health colleagues have been under enormous strain throughout 2020 dealing with the ongoing pandemic. In hospitals, particularly those with small teams, the same core group of staff would traditionally deal with AMR response, stewardship and IPC activities and the added demands of COVID-19 disproportionately affects the capacity of those teams to deal with AMR and stewardship. The increased focus on environmental hygiene throughout the pandemic has likely impacted positively on cleaning. Laboratory scientific staffing resources are already very stretched and the added demands of COVID-19 pandemic on staffing has made it even more challenging to recruit [sic]." |
| **AMR data information systems** | Low | "The biggest problem is that data are not generated as before and with the special focus on covid, dissemination platforms for data are not available and people are not paying attention to other data sets, but only covid." |
|  | Lower-middle | "We have a National AMR database where AMR data are stored, so no changes were experienced [sic]." |
|  | Upper-middle | "Hospital administration initiated planning to prevent delayed reporting [sic]." |
|  | High | "A laboratory-based surveillance system, originally implemented for AMR surveillance, was adapted to also capture data on SARS-CoV-2 testing and allow for the analysis of co-infections [sic]." |
| **Patient-case mix** | Low | "Non-urgent hospital visits and elective surgeries decreased due to the COVID-19 scare...Hospital bed occupancy and intensive care unit admission moderately increased due to COVID-19 positive cases being held for two weeks under observation. On the other end, chronically ill cases were avoiding contact with the COVID-19 situations in the hospitals [sic]." |
|  | Lower-middle | "Reduction or even stopping non-emergency hospital activities (non-urgent and elective surgical procedures) during confinement. Number of ICU beds increased in some hospitals." |
|  | Upper-middle | "We have reorganized health services. Some started to serve only COVID-19...In addition, the understanding at the beginning of the pandemic that you should only take care in case of breathing difficulties generated a low demand for emergency care." |
|  | High | "Preventive measures have been taken to reduce COVID-19 transmission such as the diversion to virtual clinics and phone consultation mainly for outpatients, delivery of medicine to homes, reducing the stay in the hospital and discharging of the patients if the clinical condition is ok, postponing the elective surgeries and working mainly on the emergency procedures and surgeries [sic]." |
| **Infection prevention and control practices** | Low | "All people and at every work station were observing hand hygiene, social distancing, alcohol hand rub, and mask wearing which positively controls spread of antimicrobial resistant organisms [sic]." |
|  | Lower-middle | "Our various hospital structures took advantage of this situation to strengthen their IPC activities (particularly awareness, training)." |
|  | Upper-middle | "Several campaigns have been held including WASH awareness campaigns." |
|  |  | "Training is not possible due to strict social distancing. Virtual meetings are not practical if the IT system is not well supported [sic]." |
|  | High | "IPC staff was overworked by COVID-19 and IPC training was performed by peers (by peers and IPC link nurses)."<br>"COVID-19 has revealed the need to integrate infection prevention and control across the entire healthcare delivery system. This needed response includes strategies for implementation across all levels of care, use of data for targeted action, tailored tools and strategies for early detection and management, effective ongoing communication and education, strong connection between public health and healthcare, policies for accountability and sustainability, and an ongoing commitment to these improvements." |
| **Antibiotic consumption** | Low | "Due to the fever and other presenting symptoms of COVID-19, patients try to do self-medication and doctors also prescribe antibiotics empirically since the infecting agent was not able to be cultured then." |
|  | Lower-middle | "Consumption of WHO watch and reserve antibiotics increased because in rural facilities where diagnostic tools for COVID-19 are scarce, the use of antibiotics for pneumonia-like symptoms increased and in urban facilities, the use of azithromycin is still frequent [sic]." |
|  | Upper-middle | "For large hospitals, there was no impact. For smaller hospitals, moderate decrease was observed due to decreasing patient numbers [sic]." |
|  | High | "Preliminary data show slight increase in March/April in watch antibiotics (such as azithromycin/carbapenems) in inpatient settings [sic]."<br>"We will look at this in more detail, but good data are not yet available." |
| **Antimicrobial resistance rates** | Low | "Each infection with pathogens avoidable by hygiene for us decreased." |
|  | Lower-middle | "It's too early to comment on impact of increase in use of antimicrobials on AMR during the pandemic, we may have better idea of the impact on AMR trends over the next couple of years." |
|  | Upper-middle | "For large hospitals, there was no impact. For smaller, a moderate decrease was observed due to decreasing patient number [sic]." |
|  | High | "The national 2020 antibiotic resistance data will be available only next year, so it is too early to assess the impact of COVID-19 on AMR rates. However, the impression is that MDR isolates are more frequent in ICUs caring for COVID patients. Also, decreased sampling in COVID ICUs, due to the lack of personnel may contribute to underestimating the problem of MDR." |
| **Long-term perspectives** | Low | "If no concerted work is not done to control the spread of AMR it will be another pandemic to hit the world." |
|  | Lower-middle | "We need to balance AMR and COVID response activities." |
|  | Upper-middle | "We need to harness the potential of virtual and remote working methods, this will allow us to reach a larger audience with regard to training of stewardship committee members and health facility workers in their management and surveillance of AMR." |
|  | High | "We need to support greater resiliency in antibiotic resistance and antibiotic use programs in healthcare and public health. Because, without this resiliency, critical AR work will not happen as new threats emerge."<br>"It is important that pauses in AMR and HCAI surveillance and stewardship activities are short-term and that experienced staff are supported to resume these activities through recruitment of additional staff for pandemic-related work and investment in infrastructures that facilitate efficient ways of working and acknowledge remote working requirements, e.g. electronic prescribing, surveillance systems." |

**Figure 1.**
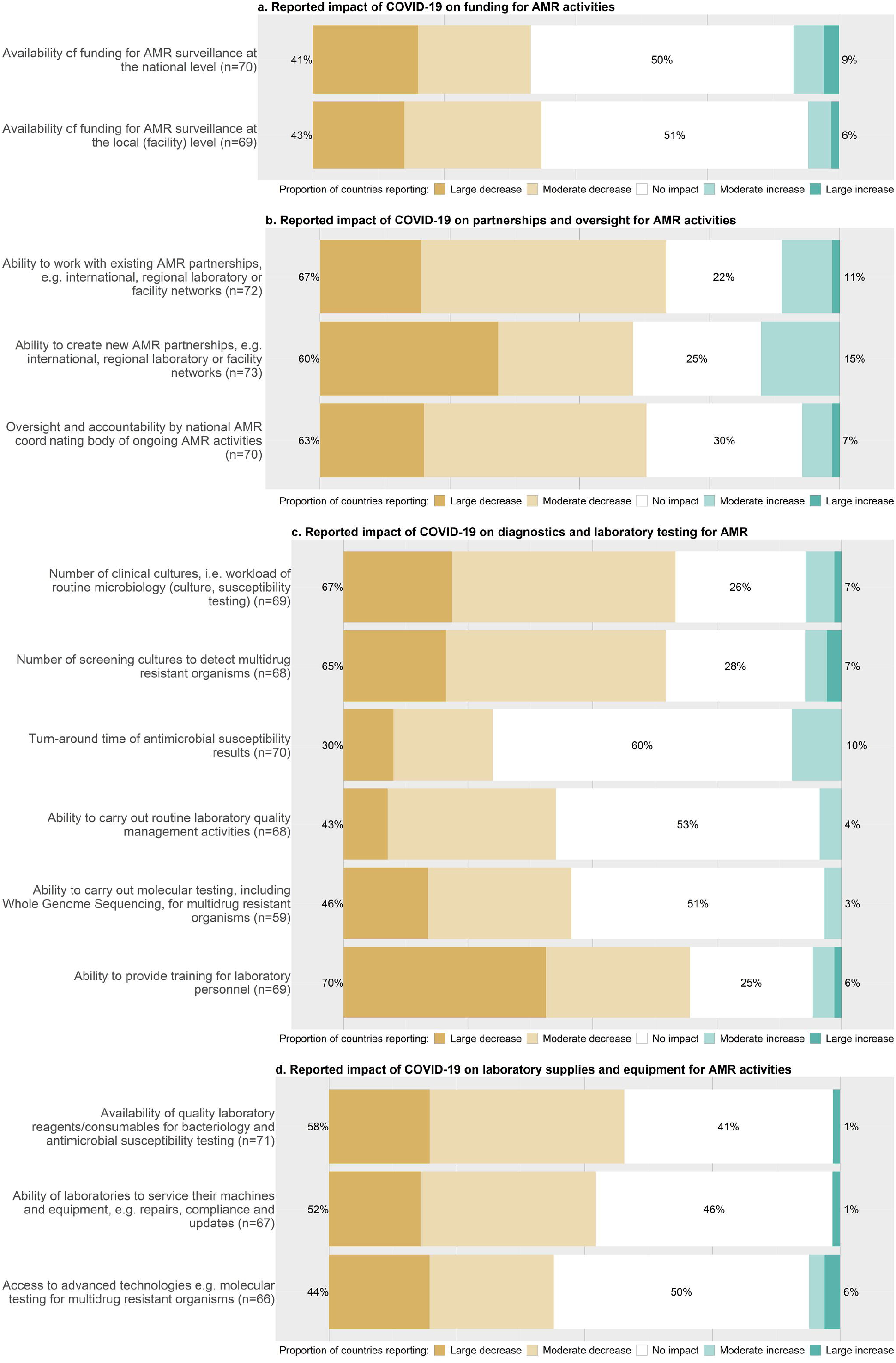
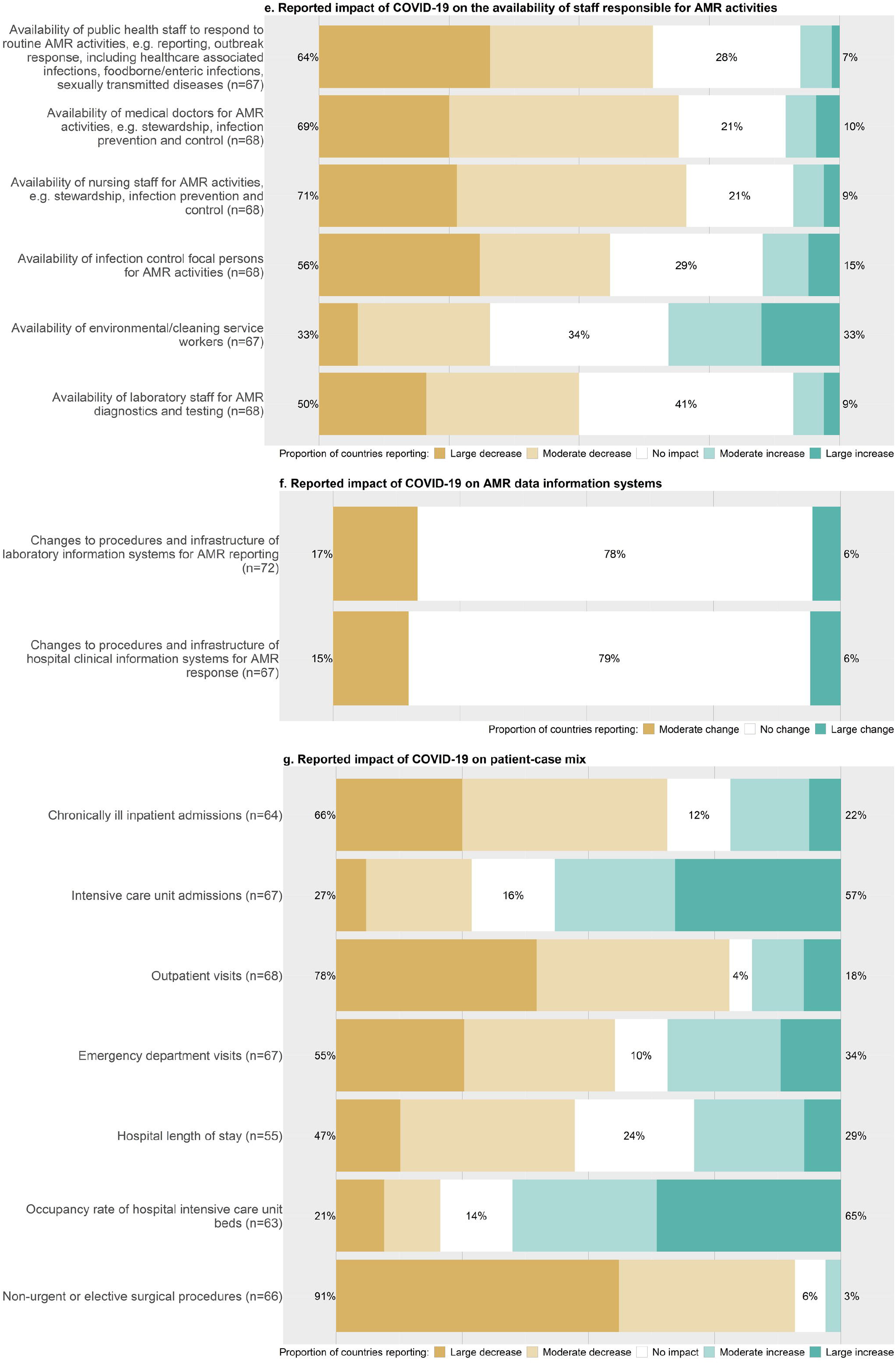
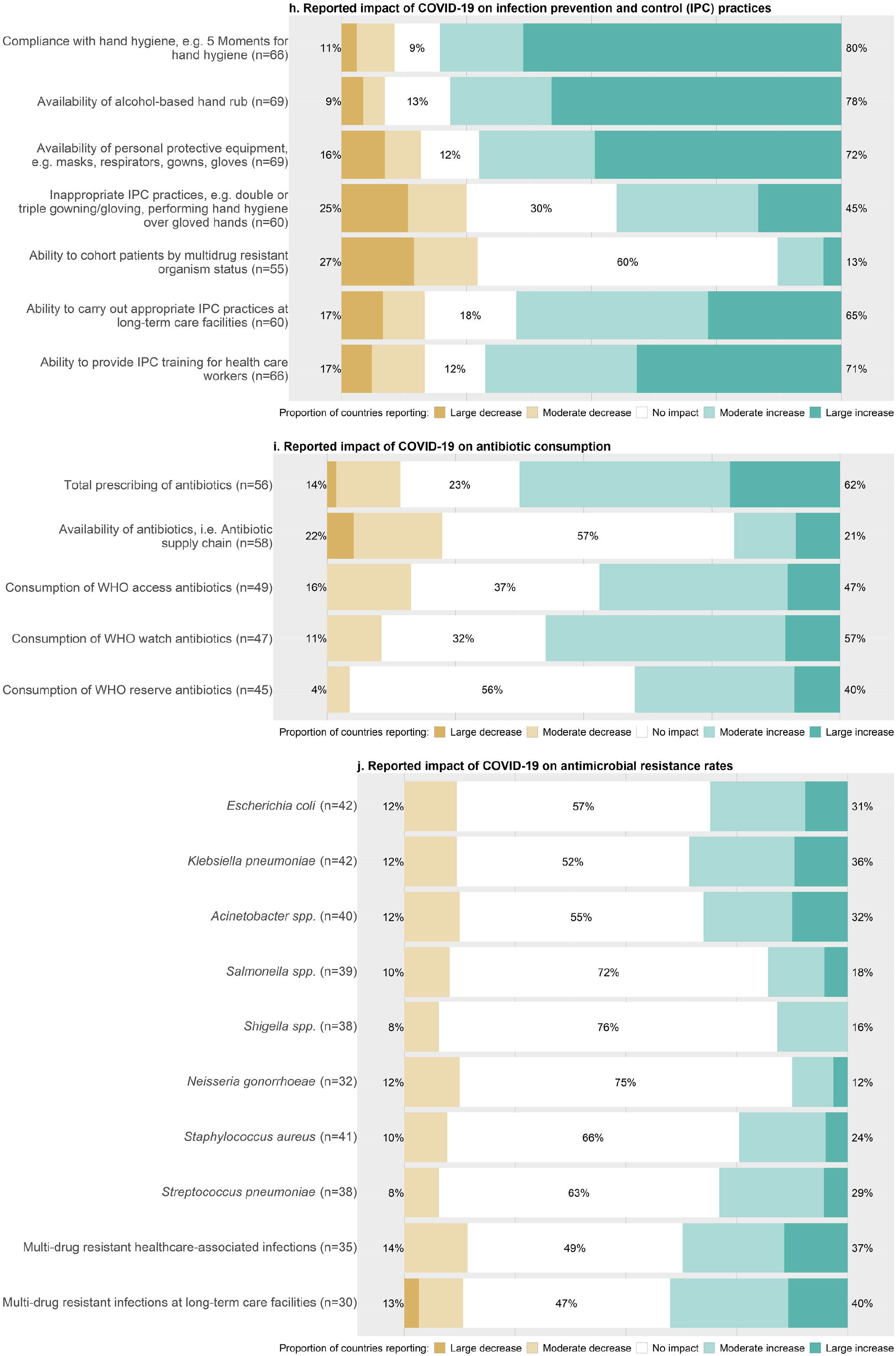
Likert responses for the impacts of COVID-19 on AMR in GLASS countries (N=73)* *Only completed responses are shown and respective denominators are shown on the left axis per question.

### Partnerships and oversight for AMR activities

More than half of responding countries reported decreases in their ability to work with new (44/73; 60%) or existing (48/72; 67%) partnerships, as well as in the ability of the national AMR coordinating body to have oversight of activities (44/70; 63%) (Figure 1b). In contrast, eleven (11/73; 15%) countries reported increases in the ability to create new partnerships since the COVID-19 pandemic. No significant income level or regional differences were seen. In the free-text questions (Table 2), a few low- and middle-income countries (LMICs) reported compounding challenges affecting partnerships, such as mobility restrictions and poor internet connections. Selected middle- and high-income countries described opportunities such as the creation of new partnership platforms, possibilities for data exchange, and identified gaps relevant for both COVID-19 and AMR action-planning.

### Diagnostics and laboratory testing for AMR

A majority of countries responding reported decreases in the number of screening (65%; 44/68) and clinical (67%; 46/69) cultures requested (Figure 1c). Although more than half of countries (42/70; 60%) reported no impact on the turn-around time of antimicrobial susceptibility results (AST), many (48/69; 70%) reported decreases in their ability to provide training for laboratory personnel; 46% (27/59) and 43% (29/68) also reported decreases in the ability to carry out molecular testing and quality management activities, respectively. No significant income level or regional differences were seen. In the free-text questions (Table 2), countries across income levels described influencing factors such as fewer patients visiting hospitals and the need to divert staff, equipment and reagents for COVID-19 testing. In contrast, one low-middle-income country reported that the laboratory network was strengthened in their country and expected this to have a positive impact on the response against AMR. Selected countries across income levels also highlighted the potential for leveraging COVID-19 work for AMR, such as in the area of microbial genomics and rapid testing for co-infections or secondary bacterial infections.

### Laboratory supplies and equipment for AMR activities

More than half of responding countries reported decreases in the availability of quality laboratory reagents and consumables for bacteriology and AST (41/71; 58%) and in the ability to service machines and equipment (35/67; 52%) (Figure 1d). In contrast, 41% (n=29) and 50% (n=33) of countries reported no impact on reagent/consumable availability and access to advanced technologies, respectively. Decreases in reagent/consumable availability were reported more frequently by low-(11/11) and middle-income (17/31) countries compared to high-income (11/26) countries, respectively (p<0.01). Decreases were also more frequently reported by the African and Eastern Mediterranean regions compared to other regions (p<0.01). In the free-text questions (Table 2), countries reported broad difficulties receiving supplies due to travel and import restrictions. High-income countries reported more specific impacts on particular supplies due to COVID-19 testing, such as the availability of molecular diagnostic platforms.

### Availability of staff responsible for AMR activities

Responding countries reported the largest decreases in the availability of nursing (48/68; 71%), medical (47/68; 69%) and public health staff (43/67; 64%) for AMR activities such as stewardship, outbreak response and reporting (Figure 1e). In contrast, 67% of countries (45/67) reported no impact or increases in the availability of environmental cleaning workers. Increases in the availability of IPC focal points were reported by 56% (38/68) and more frequently by low-(4/11) and middle-income (5/32) countries compared to high-income (0/22) countries (p=0.03). In the free-text questions (Table 2), many countries across income levels described significant public health, medical, nursing and laboratory workforce challenges. One high-income country stated that there had been a reduction in staff due to closed borders. Selected countries across income levels highlighted the importance of aligning COVID-19 and AMR staff trainings. One middle-income country requested increased virtual communication between GLASS national focal points on AMR in the context of COVID-19.

### AMR data information systems

Most countries who responded reported no impact on clinical (53/67; 79%) or laboratory (56/72; 78%) data information systems for AMR (Figure 1f). No significant income level or regional differences. In the free-text questions (Table 2), one low-income country reported decreases in the use of AMR data information systems during the pandemic, whereas one high-income country highlighted potential opportunities for integrating AMR and SARS-CoV-2 data.

### Patient-case mix

Most responding countries reported decreases in the number of elective surgical procedures (60/66; 91%), outpatient visits (53/68; 78%) and chronically ill inpatient admissions (42/64; 66%), while more than half of countries reported increases in intensive care unit (ICU) admissions (38/67; 57%) and the occupancy rate of ICU beds (41/63; 65%) (Figure 1g). Decreases in outpatient visits were reported more frequently by high-(21/24) and middle-income (24/30) countries compared to low-income (6/11) countries (p=0.04). In the free-text questions (Table 2), countries across income levels reported changes due to the reorganisation of healthcare services such as hospitals prioritising only COVID-19 patients, reducing non-emergency services, and diversion to online or phone consultations. A small number of LMICs reported the closure of primary health care facilities for several months as well as the perception that patients were avoiding all health care facilities during the pandemic.

### IPC practices

Many responding countries reported improvements in IPC as a result of COVID-19. This included a majority reporting increases in hand hygiene compliance (53/66; 80%), availability of alcohol-based hand rub (54/69; 78%), availability of personal protective equipment (PPE; 50/69; 72%), and ability to carry out IPC training (47/66; 71%) (Figure 1h). More than half of countries reported no impact on the ability to cohort patients by multidrug-resistant organism status (33/55, 60%). In contrast, 45% (27/60) of countries reported an increase in inappropriate IPC practices such as double gowning or gloving and performing hand hygiene over gloved hands. No significant income level or regional differences were seen. In the free-text questions (Table 2), countries of all income levels highlighted the strengthening of IPC efforts through awareness campaigns and training. However, one middle-income country reported social distancing and poor internet connection as barriers to delivering training and one high-income country reported the use of peers and link nurses for training due to overworked IPC staff. Several middle-and high-income countries also highlighted IPC lessons learned from COVID-19 such as the need to better integrate IPC across the entire healthcare delivery system and the need for earlier focus on IPC implementation in long-term care facilities.

### Antibiotic consumption

More than half of responding countries (35/56; 63%) reported increases in total prescribing of antibiotics (Figure 1i). More specifically, 47% (23/49), 57% (27/47) and 40% (18/45) of countries reported increased use of WHO Access, Watch, and Reserve antibiotics, respectively. More than half of countries (33/58; 57%) reported no impact on the availability of antibiotics. Increases in total prescribing were reported more frequently by low-(8/10) and middle-income (18/24) countries compared to high-income (7/20) countries (p=0.03). In the free-text questions (Table 2), countries across income levels highlighted preliminary data suggesting increases in antibiotic use although many were also not yet able to assess. Selected high-income countries specified increases in the use of watch and reserve antibiotics such as azithromycin at health care facilities. One middle-income country reported that antibiotics are being prescribed in almost all cases of COVID-19 regardless of indications and one low-income country reported that more people in the community were self-prescribing antibiotics. In contrast, a few middle- and high-income countries reported reductions in levels of prescribing due to less healthcare utilization.

### AMR rates

Fewer countries were able to report on the impacts of COVID-19 on AMR rates. Among those that responded, 37% (13/35) and 40% (12/30) of countries reported increases in multi-drug resistant (MDR) healthcare-associated infections and MDR infections in long-term care facilities, respectively (Figure 1j). Approximately one third of countries reported increases in Gram negative organisms such as *Klebsiella pneumoniae* (15/42; 36%), *Acinetobacter* spp. (13/40; 32%), and *Escherichia coli* (13/42; 31%) and Gram-positive organisms such as *Staphylococcus aureus* (24%; 10/41) and *Streptococcus pneumoniae* (29%; 11/38). No impacts on selected organisms were reported more frequently by high- and middle-income countries compared to low-income countries (p<0.01). In the free-text questions (Table 2), many countries across income levels reported that they were not yet able to reliably report on AMR data. Several high-income countries suggested that resistance rates may be higher as a result of more patients being treated in ICUs or long-term care settings. A few LMICs suggested that there may be a reduction in resistance due to fewer patients presenting to the hospital overall.

### Long-term perspectives

In the free-text questions (Table 2), predictions from countries across income levels on the long-term impacts of the pandemic on AMR were mixed, citing factors that could reduce resistance, such as improved IPC, versus factors that could increase resistance, such as worsening of antimicrobial stewardship practices, increased staff fatigue to detect AMR threats, and reduced prioritization of AMR initiatives in place of those on COVID-19. One middle-income country suggested that the negative effects on AMR were greatest at the start of the pandemic. Many countries across income levels highlighted the need to balance COVID-19 and AMR response activities and address gaps in funding, staffing, consumables, equipment and IT infrastructure. A few middle- and high-income countries suggested the need to develop tools such as improved guidelines on antibiotic prescribing for COVID-19 patients and expanded rapid and molecular testing. A few countries across income levels emphasized the importance of continued health system strengthening and resiliency including IPC and AMR awareness.

## Discussion

The COVID-19 pandemic is having wide-reaching impacts on all aspects of our health systems. These have upended various levels of AMR surveillance, prevention and control. Including a wide range of country settings, this survey gives an important initial picture of the global impacts that COVID-19 has on these AMR aspects. Responses from GLASS national focal points revealed some universal patterns but also captured the variability across countries which, in some cases, could be linked to income level. The reported impacts involved factors that could bias AMR reporting as well as potentially decrease or increase AMR rates. These findings provide a useful framework to inform the ongoing implementation of AMR surveillance and interpretation of data. They also present key country insights on how we might use the COVID-19 response to make continued gains in combatting AMR.

Reported aspects which could bias AMR data included a range of changes in health care utilisation, testing activities and diagnostic resources. These included decreases in the number of cultures, elective surgeries, chronically ill admissions and outpatient visits (particularly in LMICs) as well as increases in ICU admissions. Other observational and quasi-experimental studies have similarly documented reductions in global surgical case volume proxies as well as hospital emergency department visits particularly in high-income countries.^10-12^ Such changes in patients and testing denominators should be considered where feasible when interpreting changes in AMR data and potential biases.

Many countries reported IPC improvements which could favour the prevention of both AMR and COVID-19, such as hand hygiene, PPE use, the stable availability of environmental cleaning workers and increased availability of IPC focal points (particularly in LMICs). WHO has highlighted the importance of effective COVID-19 IPC measures including compliance with standard and transmission-based precautions,^13^ and many countries and facilities seem to be taking steps to promote this guidance. Improved IPC awareness and implementation has been shown to improve after large outbreaks, as also seen after the 2014/2016 Ebola outbreak.^14^ It is an opportunity that can be utilised to promote sustainable IPC programmes that can more effectively combat emerging threats such as COVID-19 and Ebola as well as AMR transmission.

Increased total antibiotic prescribing was reported by more than half of countries, particularly by LMICs. Specific increases in WHO Watch antibiotics (e.g. azithromycin) were also seen despite conflicting evidence for COVID-19 patients.^15^ Meta-analyses by Langford et al. and Rawson et al. found that approximately three quarters of hospitalized COVID-19 patients received antibiotics, although 3.5% and 8.5% were estimated to have bacterial co-infections on presentation and bacterial/fungal co-infections during admission, respectively.^16, 17^ Hospital-based studies have shown significant increases in total antibiotic use, including broad-spectrum antibiotic use such as cefepime, piperacillin/tazobactam and carbapenems.^18-20^ National outpatient studies have found both decreases in antibiotic prescriptions due to COVID-19 restrictions^21, 22^ and increases in expected prescriptions after controlling for changes in the number of telephone consultations versus in-person appointments.^23^ In light of these results, evidence-based guidelines such as the indications for antibacterial therapy in COVID-19 patients should be promoted globally to prevent the accelerated threat of AMR.^24-26^

Although improved IPC efforts should be leveraged for both COVID-19 and AMR, an overly simplistic approach should also be avoided. Not all IPC measures for respiratory disease will be effective against AMR transmission. Inappropriate IPC practices (e.g. double gloving) reported by half of countries in this survey should be avoided. Other studies have also reported the unintended consequences of inappropriate PPE practices during the COVID-19 and SARS outbreaks, leading to higher risks of contamination.^27-29^

Various programmatic and structural factors which could reduce AMR activities were also reported by countries. Decreases in the routine ability to work with partnerships, funding, staffing and supplies could lead to important gaps in communication, implementation and data exchange. Specific challenges related to border closures, blocked imports and competition for limited stocks of material leading to disparities across countries have been highlighted in other anecdotal reports.^3-6, 30, 31^ Despite these challenges, some countries reported efforts to identify areas where the COVID-19 and AMR responses overlap (e.g. integrated stewardship guidance, new partnership platforms, leverage of funding for health systems strengthening and overall laboratory network improvements). These strategies are critical to maintaining routine AMR control alongside the COVID-19 response. It is equally important to recognise where response activities are not the same and additional efforts are needed, such as investments in staffing and back-up suppliers.

Most countries did not yet have complete data on AMR rates. It is still early in the course of the pandemic to reliably report on any changes. Although it will take time to effectively analyse these data, reported factors such as decreases in surveillance capacity could limit the ability to provide data on true AMR changes. It is critical that AMR activities remain a priority for countries and high on the global health agenda to ensure the necessary capacity to detect and respond to emerging threats. Outbreaks and increases in AMR acquisition, such as carbapenemase-producing Enterobacterales, during the COVID-19 pandemic have been reported by hospitals, demonstrating the importance of continuing routine AMR activities and closely monitoring these data.^19, 32-35^

Several survey limitations should be considered. Although these results provide a useful initial global snapshot of the impacts of COVID-19 on AMR, they could also be considered an oversimplification of complex and varying experiences across national and facility levels. Heterogeneity between countries in the robustness of their existing AMR surveillance systems and activities (i.e. capacity, experience, resources) may have affected survey interpretation. If national focal points did not coordinate their responses with other experts or did not have sufficient knowledge of the data or dynamics in their country, the validity and reliability of some responses may have been affected. Furthermore, self-reported perceptions of national focal points on the impacts of COVID-19 on AMR could have also been affected by self-desirability bias.

This survey was the first concerted effort to explore the global impact of COVID-19 on AMR surveillance, prevention and control among GLASS countries. Although much focus is understandably placed on COVID-19 at present, we believe that AMR must remain high on the global health agenda. This survey included a wide range of country settings and responses which highlighted both the universal effects of COVID-19 on AMR as well as the heterogeneity of impacts across countries. The results underscore the importance of finding ways to leverage the COVID-19 response activities to also support routine AMR prevention and control, where possible, and advocate for continued investments in IPC and laboratory strengthening for overall health system preparedness. It is critical to continue to monitor the dynamic situation and update national action plans with lessons learned to include preparedness and mitigation for future emerging threats that may also affect routine AMR work. Countries are encouraged to engage with GLASS and the Network to work collectively towards leveraging opportunities and addressing present challenges to improve AMR surveillance, prevention and control.

## Data Availability

The aggregated data is avaiable on a case-by-case basis upon consultation with the WHO AMR Surveillance and Quality Assessment Collaborating Centres Network coordinator (Robert Koch Institute, Berlin, Germany)

## Acknowledgements

We would like to acknowledge all of the WHO Collaborating Centres that are part of the WHO AMR Surveillance and Quality Assessment Collaborating Centres Network: https://www.who.int/glass/collaborating-centres-network/en/. In addition, we would like to acknowledge Muna Abu Sin and Birgitta Schweickert (Robert Koch Institute, Berlin, Germany) who provided feedback on the questionnaire structure. We would like to sincerely thank all of the participating countries enrolled in GLASS at the time of the survey. Those that agreed to be acknowledged included: Afghanistan, Argentina, Australia, Bahrain, Bosnia and Herzegovina, Croatia, Djibouti, Finland, France, Gabon, Gambia, Georgia, Germany, Greece, India, Indonesia, Iraq, Ireland, Japan, Kenya, Lebanon, Liberia, Lithuania, Madagascar, Mali, Morocco, Namibia, Netherlands, Oman, Pakistan, Palestine, Poland, Qatar, Russian Federation, Singapore, South Africa, Sudan, Switzerland, Syrian Arab Republic, Thailand, Trinidad and Tobago, Tunisia, Uganda, United Arab Emirates, United Republic of Tanzania, United States of America, and Zambia.

## Transparency

None to declare.

## Disclaimer

The findings and conclusions in this report are those of the authors and do not necessarily represent the official positions of their affiliated institutions.

## Supplementary data

Supplementary Materials are available at the end of this manuscript.

## References

1. World Health Organization. Global Action Plan on Antimicrobial Resistance. https://www.who.int/antimicrobial-resistance/global-action-plan/en/.

2. World Health Organization. Record number of countries contribute data revealing disturbing rates of antimicrobial resistance. https://www.who.int/news/item/01-06-2020-record-number-of-countries-contribute-data-revealing-disturbing-rates-of-antimicrobial-resistance.

3. Getahun H, Smith I, Trivedi K et al. Tackling antimicrobial resistance in the COVID-19 pandemic. Bull World Health Organ 2020; 98: 442–a.

4. Donà D, Di Chiara C, Sharland M. Multi-drug-resistant infections in the COVID-19 era: a framework for considering the potential impact. J Hosp Infect 2020; 106: 198–9.

5. Monnet DL, Harbarth S. Will coronavirus disease (COVID-19) have an impact on antimicrobial resistance? Euro Surveill 2020; 25: 2001886.

6. Chibabhai V, Duse AG, Perovic O et al. Collateral damage of the COVID-19 pandemic: Exacerbation of antimicrobial resistance and disruptions to antimicrobial stewardship programmes? S Afr Med J 2020; 110: 572–3.

7. World Health Organization. WHO AMR Surveillance and Quality Assessment Collaborating Centres Network. https://www.who.int/glass/collaborating-centres-network/en/.

8. World Health Organization. Monitoring the building blocks of health systems. https://www.who.int/healthinfo/systems/WHO_MBHSS_2010_full_web.pdf?ua=1.

9. World Bank. The world by income. https://datatopics.worldbank.org/world-development-indicators/the-world-by-income-and-region.html.

10. O’Reilly-Shah Vn, Van Cleve W, Long DR et al. Impact of COVID-19 response on global surgical volumes: an ongoing observational study. Bull World Health Organ 2020; 98: 671–82.

11. Mulholland RH, Wood R, Stagg HR et al. Impact of COVID-19 on accident and emergency attendances and emergency and planned hospital admissions in Scotland: an interrupted time-series analysis. J R Soc Med 2020; 113: 444–53.

12. Sokolski M, Gajewski P, Zymlinski R et al. Impact of Coronavirus Disease 2019 (COVID-19) Outbreak on Acute Admissions at the Emergency and Cardiology Departments Across Europe. Am J Med.

13. World Health Organization. Infection prevention and control during health care when coronavirus disease (COVID-19) is suspected or confirmed. https://www.who.int/publications/i/item/WHO-2019-nCoV-IPC-2020.4.

14. Tremblay N, Musa E, Cooper C et al. Infection prevention and control in health facilities in post-Ebola Liberia: don’t forget the private sector! Public Health Action 2017; 7: S94–s9.

15. Verdejo C, Vergara-Merino L, Meza N et al. Macrolides for the treatment of COVID-19: a living, systematic review. Medwave 2020; 20: e8074.

16. Langford BJ, So M, Raybardhan S et al. Antibiotic prescribing in patients with COVID-19: rapid review and meta-analysis. Clin Microbiol Infect.

17. Rawson TM, Moore LSP, Zhu N et al. Bacterial and Fungal Coinfection in Individuals With Coronavirus: A Rapid Review To Support COVID-19 Antimicrobial Prescribing. Clin Infect Dis 2020.

18. Abelenda-Alonso G, Padullés A, Rombauts A et al. Antibiotic prescription during the COVID-19 pandemic: A biphasic pattern. Infect Control Hosp Epidemiol 2020; 41: 1371–2.

19. Bork JT, Leekha S, Claeys K et al. Change in hospital antibiotic use and acquisition of multidrug-resistant gram-negative organisms after the onset of coronavirus disease 2019. Infect Control Hosp Epidemiol 2020: 1–3.

20. Gonzalez-Zorn B. Antibiotic use in the COVID-19 crisis in Spain. Clin Microbiol Infect.

21. King LM, Lovegrove MC, Shehab N et al. Trends in U.S. outpatient antibiotic prescriptions during the COVID-19 pandemic. Clin Infect Dis 2020.

22. Orubu ESF, Malik F, Figueras A et al. Antibacterial consumption in the context of COVID-19 in Pakistan: an analysis of national pharmaceutical sales data for 2019-20. medRxiv 2020: 2020.12.05.20244657.

23. Armitage R, Nellums LB. Antibiotic prescribing in general practice during COVID-19. Lancet Infect Dis.

24. Sieswerda E, de Boer Mgj, Bonten MMJ et al. Recommendations for antibacterial therapy in adults with COVID-19 -an evidence based guideline. Clin Microbiol Infect 2021; 27: 61–6.

25. National Institute for Health and Care Excellence (UK). COVID-19 rapid guideline: antibiotics for pneumonia in adults in hospital. https://www.ncbi.nlm.nih.gov/books/NBK566162/.

26. World Health Organization. COVID-19 Clinical management: living guidance. https://www.who.int/publications/i/item/WHO-2019-nCoV-clinical-2021-1.

27. Yap FHY, Gomersall CD, Fung KSC et al. Increase in Methicillin-Resistant Staphylococcus aureus Acquisition Rate and Change in Pathogen Pattern Associated with an Outbreak of Severe Acute Respiratory Syndrome. Clin Infect Dis 2004; 39: 511–6.

28. Meda M, Gentry V, Reidy P et al. Unintended consequences of long-sleeved gowns in a critical care setting during the COVID-19 pandemic. J Hosp Infect 2020; 106: 605–9.

29. Stevens MP, Doll M, Pryor R et al. Impact of COVID-19 on traditional healthcare-associated infection prevention efforts. Infect Control Hosp Epidemiol 2020; 41: 946–7.

30. Hsu J. How covid-19 is accelerating the threat of antimicrobial resistance. Br Med J 2020; 369: m1983.

31. Lucien MAB, Canarie MF, Kilgore PE et al. Antibiotics and antimicrobial resistance in the COVID-19 era: Perspective from resource-limited settings. Int J Infect Dis 2021; 104: 250–4.

32. Tiri B, Sensi E, Marsiliani V et al. Antimicrobial Stewardship Program, COVID-19, and Infection Control: Spread of Carbapenem-Resistant Klebsiella Pneumoniae Colonization in ICU COVID-19 Patients. What Did Not Work? J Clin Med 2020; 9.

33. Cantón R, Akóva M, Carmeli Y et al. Rapid evolution and spread of carbapenemases among Enterobacteriaceae in Europe. Clin Microbiol Infect 2012; 18: 413–31.

34. Farfour E, Lecuru M, Dortet L et al. Carbapenemase-producing Enterobacterales outbreak: Another dark side of COVID-19. Am J Infect Control 2020; 48: 1533–6.

35. Gomez-Simmonds A, Annavajhala MK, McConville TH et al. Carbapenemase-producing Enterobacterales causing secondary infections during the COVID-19 crisis at a New York City hospital. J Antimicrob Chemother 2021; 76: 380–4.

