## Supplementary data for "Impact of the COVID-19 Pandemic on Antimicrobial Resistance (AMR) Surveillance, Prevention and Control: A Global Survey"

**Appendix 1. Questionnaire**

| **Which country are you responding on behalf of?** | [Drop-down list with countries, including “prefer not to respond”] *Official GLASS country list publically available here, and categorised by income level:* |
|---|---|

*Please indicate your **<u>perceptions</u>** of the impact of COVID-19 (i.e. from large decrease to large increase) in the following areas since the start of the COVID-19 epidemic to date in your country.*

| **1. Do you think there has been an impact on <u>funding for AMR activities</u> as a result of COVID-19 in your country in terms of the following:** | Large decrease | Moderate decrease | No impact | Moderate increase | Large increase | Do not know |
|---|---|---|---|---|---|---|
| Availability of funding for AMR surveillance at the national level | | | | | | |
| Availability of funding for AMR surveillance at the local (facility) level | | | | | | |
| ***Please provide any further information you may have in this area (e.g. explaining a particular increase/decrease, examples/data):*** Free text response | | | | | | |

| **2. Do you think there has been an impact on <u>partnerships and oversight for AMR activities</u> as a result of COVID-19 in your country in terms of the following:** | Large decrease | Moderate decrease | No impact | Moderate increase | Large increase | Do not know |
|---|---|---|---|---|---|---|
| Ability to work with existing AMR partnerships, e.g. international, regional laboratory or facility networks | | | | | | |
| Ability to create new AMR partnerships, e.g. international, regional laboratory or facility networks | | | | | | |
| Oversight and accountability by national AMR coordinating body of ongoing AMR activities | | | | | | |
| ***Please provide any further information you may have in this area (e.g. explaining a particular increase/decrease, examples/data):*** Free text response | | | | | | |

| **3. Do you think there has been an impact on <u>diagnostics and laboratory testing for AMR</u> as a result of COVID-19 in your country in terms of the following:** | Large decrease | Moderate decrease | No impact | Moderate increase | Large increase | Do not know |
|---|---|---|---|---|---|---|
| Number of clinical cultures, i.e. workload of routine microbiology (culture, susceptibility testing) | | | | | | |
| Number of screening cultures to detect multidrug resistant organisms | | | | | | |
| Turn-around time of antimicrobial susceptibility results | | | | | | |
| Ability to carry out routine laboratory quality management activities | | | | | | |
| Ability to carry out molecular testing, including Whole Genome Sequencing, for multidrug resistant organisms | | | | | | |
| Ability to provide training for laboratory personnel | | | | | | |

| |
|---|
| ***Please provide any further information you may have in this area (e.g. explaining a particular increase/decrease, examples/data):*** Free text response |

| **4. Do you think there has been an impact on <u>laboratory supplies and equipment for AMR activities</u> as a result of COVID-19 in your country in terms of the following:** | Large decrease | Moderate decrease | No impact | Moderate increase | Large increase | Do not know |
|---|---|---|---|---|---|---|
| Availability of quality laboratory reagents/consumables for bacteriology and antimicrobial susceptibility testing | | | | | | |
| Ability of laboratories to service their machines and equipment, e.g. repairs, compliance and updates | | | | | | |
| Access to advanced technologies e.g. molecular testing for multidrug resistant organisms | | | | | | |
| ***Please provide any further information you may have in this area (e.g. explaining a particular increase/decrease, examples/data):*** Free text response | | | | | | |

| **5. Do you think there has been an impact on the <u>availability of staff responsible for AMR activities</u> as a result of COVID-19 in your country in terms of the following:** | Large decrease | Moderate decrease | No impact | Moderate increase | Large increase | Do not know |
|---|---|---|---|---|---|---|
| Availability of public health staff to respond to routine AMR activities, e.g. reporting, outbreak response, including healthcare associated infections, foodborne/enteric infections, sexually transmitted diseases | | | | | | |
| Availability of medical doctors for AMR activities, e.g. stewardship, infection prevention and control | | | | | | |
| Availability of nursing staff for AMR activities, e.g. stewardship, infection prevention and control | | | | | | |
| Availability of infection control focal persons for AMR activities | | | | | | |
| Availability of environmental/cleaning service workers | | | | | | |
| Availability of laboratory staff for AMR diagnostics and testing | | | | | | |
| ***Please provide any further information you may have in this area (e.g. explaining a particular increase/decrease, examples/data):*** Free text response | | | | | | |

| **6. Do you think there has been an impact on <u>AMR data information systems</u> as a result of COVID-19 in your country in terms of the following:** | Large change | Moderate change | No change | Do not know |
|---|---|---|---|---|
| Changes to procedures and infrastructure of *laboratory* information systems for AMR reporting | | | | |
| Changes to procedures and infrastructure of *hospital clinical* information systems for AMR response | | | | |
| ***Please provide any further information you may have in this area (e.g. explaining a particular increase/decrease, examples/data):*** Free text response | | | | |

| **7. Do you think there has been an impact on <u>patient-case mix</u> as a result of COVID-19 in your country in terms of the following:** | Large decrease | Moderate decrease | No impact | Moderate increase | Large increase | Do not know |
|---|---|---|---|---|---|---|
| Chronically ill inpatient admissions | | | | | | |
| Intensive care unit admissions | | | | | | |
| Outpatient visits | | | | | | |
| Emergency department visits | | | | | | |
| Hospital length of stay | | | | | | |
| Occupancy rate of hospital intensive care unit beds | | | | | | |
| Non-urgent or elective surgical procedures | | | | | | |
| ***Please provide any further information you may have in this area (e.g. explaining a particular increase/decrease, examples/data):*** Free text response | | | | | | |

| **8. Do you think there has been an impact on <u>infection prevention and control (IPC) practices</u> as a result of COVID-19 in your country in terms of the following:** | Large decrease | Moderate decrease | No impact | Moderate increase | Large increase | Do not know |
|---|---|---|---|---|---|---|
| Compliance with hand hygiene, e.g. 5 Moments for hand hygiene | | | | | | |
| Availability of alcohol-based hand rub | | | | | | |
| Availability of personal protective equipment, e.g. masks, respirators, gowns, gloves | | | | | | |
| Inappropriate IPC practices e.g. double or triple gowning/gloving, performing hand hygiene over gloved hands | | | | | | |
| Ability to cohort patients by multidrug resistant organism status | | | | | | |
| Ability to carry out appropriate IPC practices at long-term care facilities | | | | | | |
| Ability to provide IPC training for health care workers | | | | | | |
| ***Please provide any further information you may have in this area (e.g. explaining a particular increase/decrease, examples/data):*** Free text response | | | | | | |

| **9. Do you think there has been an impact on <u>antibiotic consumption</u> as a result of COVID-19 in your country in terms of the following:** | Large decrease | Moderate decrease | No impact | Moderate increase | Large increase | Do not know |
|---|---|---|---|---|---|---|
| Total prescribing of antibiotics | | | | | | |
| Availability of antibiotics, i.e. Antibiotic supply chain | | | | | | |
| Consumption of WHO *access* antibiotics, i.e. first- or second-line treatment for common infections | | | | | | |

| |
|---|
| Consumption of WHO *watch* antibiotics, i.e. treatment for limited group of syndromes, should be monitored |
| Consumption of WHO *reserve* antibiotics, i.e. "last resort" to treat multi- or extensively-drug resistant bacteria |
| ***Please provide any further information you may have in this area (e.g. explaining a particular increase/decrease, examples/data):*** Free text response |

| **10. Do you think there has been an impact on antimicrobial resistance rates as a result of COVID-19 in your country in terms of the following:** | Large decrease | Moderate decrease | No impact | Moderate increase | Large increase | Do not know |
|---|---|---|---|---|---|---|
| *Escherichia coli* | | | | | | |
| *Klebsiella pneumoniae* | | | | | | |
| *Acinetobacter spp.* | | | | | | |
| *Staphylococcus aureus* | | | | | | |
| *Streptococcus pneumoniae* | | | | | | |
| *Salmonella* spp. | | | | | | |
| *Shigella* spp. | | | | | | |
| *Neisseria gonorrhoeae* | | | | | | |
| Multi-drug resistant healthcare-associated infections | | | | | | |
| Multi-drug resistant infections at long-term care facilities | | | | | | |
| ***Please provide any further information you may have in this area (e.g. explaining a particular increase/decrease, examples/data):*** Free text response | | | | | | |

| | |
|---|---|
| **11. Do you have any success stories you are able to share e.g. solutions to AMR challenges or opportunities to improve AMR control during the COVID-19 crisis?** | Free text response |

| | |
|---|---|
| **12. What do you predict will be the long-term impacts on AMR?** | Free text response |

| | |
|---|---|
| **13. Do you have any suggestions for improvements to the impact that the COVID-19 pandemic is having on AMR activities?** | Free text response |

| | |
|---|---|
| **14. Do you have any other comments?** | Free text response |

☐ **Our country would like to be acknowledged in any relevant publications.**

**Appendix 2. Coding framework for free-text responses from survey**

| | |
|---|---|
| Funding for AMR activities | 1. Reduction in funding for AMR activities as result of COVID-19<br>1.1. COVID-19 funding prioritised over AMR<br>1.2. Funding for AMR activities delayed due to COVID-19<br>2. Solution to funding AMR activities during COVID-19 pandemic<br>2.1. Leverage of COVID-19 for funding of AMR activities<br>3. Mixed picture/directions in terms of funding for AMR activities<br>4. No impact on funding for AMR activities<br>5. No budget or funding for AMR activities in the first place<br>6. Unable to assess whether COVID-19 has had an impact on funding for AMR activities due to lack of data<br>7. Prioritisation of COVID-19 activities, but no specific comment on funding for AMR activities |
| Partnerships and oversight for AMR activities | 8. Reduction/worsening in partnerships and oversight for AMR activities due to COVID-19<br>8.1. Networking/partnerships<br>8.2. International development/support<br>8.3. Strategy /NAP/oversight<br>9. Increase/improvement in partnerships and oversight for AMR activities due to COVID-19<br>9.1. Networking/partnerships<br>10. There was no AMR system in the first place (i.e. no partnerships and oversight for AMR activities that could be affected)<br>11. Other aspect of AMR that does not relate to partnerships or oversight<br>11.1. Reduction/worsening<br>11.1.1. Due to partners or funders shifting focus<br>11.2. Increase/improvement |
| Diagnostics and laboratory testing for AMR | 12. Reduction/delay in diagnostics and laboratory testing for AMR<br>12.1. Due to prioritisation of COVID-19<br>12.2. Due to reduced need<br>13. Increase in diagnostics and laboratory testing for AMR<br>14. Solutions in relation to diagnostics and laboratory testing for AMR<br>15. There was no AMR system in the first place (i.e. no diagnostics and laboratory testing for AMR that could be affected)<br>16. Activities relating to AMR generally affected (not specifically relating to laboratories)<br>17. Laboratory training<br>17.1. Reduction<br>17.2. Increase<br>18. Increase in general laboratory resources (link to AMR unclear) |

| | |
|---|---|
| Laboratory supplies and equipment for AMR activities | 19. Reduction/impact on laboratory supplies and equipment for AMR activities<br>19.1. Due to reliance on import/travel<br>19.2. Due to fewer patients<br>20. Increase in laboratory supplies and equipment for AMR activities<br>21. Few laboratory supplies and equipment for AMR activities available before COVID-19 pandemic<br>22. No impact on laboratory supplies and equipment for AMR activities |
| Availability of staff responsible for AMR | 23. Reduction in availability of staff responsible for AMR<br>23.1. Laboratory staff<br>23.2. Public health staff<br>23.3. Infectious disease specialists<br>23.4. Medical staff<br>23.5. Nursing staff<br>23.6. Cleaning staff<br>24. Solutions found for addressing reduced staff that are responsible for AMR<br>25. There was no AMR system in the first place (i.e. there were no staff responsible for AMR that could be affected)<br>26. Other staff-related activities<br>27. Other indirect staff-related increased<br>27.1. Increased Infection Prevention and Control (IPC) activities<br>27.2. Increased cleaning in healthcare facilities<br>28. Reduction in general AMR activities (no particular comment about staff responsible for AMR) |
| AMR data information systems | 29. Reduction in data transfer<br>30. Less (external) interest in AMR data<br>31. Innovation/solutions found for AMR data information systems<br>32. Unspecified changes to reporting<br>33. No impact on AMR data information systems as a result of COVID-19<br>34. No or barely any AMR data information systems system in place to start with (i.e. no AMR data information systems that could be affected) |
| Patient-case mix | 35. Patient-case mix affected by COVID-19<br>35.1. Increase in patients<br>35.1.1. ICU<br>35.2. Increased length of stay in hospitals<br>35.3. Reduction in patients<br>35.3.1. Non-urgent and elective care<br>35.3.2. Emergency care<br>35.3.3. General patients<br>36. Changes to respond to the patient-case mix challenge<br>36.1. Reorganisation/changes of services |

| | |
|---|---|
| | 36.2. Changes to staffing<br>37. Hospitals generally affected by COVID-19 (unspecified) |
| Infection prevention and control (IPC) | 38. IPC) practices increased/improved<br>38.1. Delivery of IPC training<br>38.2. Awareness<br>38.3. Hand hygiene compliance<br>38.4. Use of alcohol hand rub<br>38.5. Personal protective equipment<br>39. Mixed IPC picture in country<br>40. Solutions found for improving IPC practices<br>41. Challenges to/weaknesses of IPC practices<br>41.1. Ability to cohort patients<br>42. Lessons learnt in relation to IPC practices<br>43. Unknown impact of COVID-19 on IPC practices |
| Antibiotic consumption | 44. Increase in prescribing/demand of antibiotics due to COVID-19<br>45. Reduction in prescribing/demand of antibiotics due to COVID-19<br>45.1. Due to fewer patients in hospitals<br>45.2. Due to fewer people getting sick (non-COVID-19)<br>46. No impact on prescribing/demand of antibiotics due to COVID-19<br>47. Unable to assess impact of COVID-19 on antibiotic consumption<br>47.1. Due to not using use AWaRe classification<br>47.2. Due to lack of available data<br>48. Impact of COVID-19 on supply of antibiotics<br>48.1. Not known<br>48.2. Reduced |
| Antibiotic resistance | 49. Increase<br>49.1. High dependency/intensive care unit (ICU) settings<br>50. Decrease in resistance of some antibiotics/in some healthcare settings Community settings<br>50.1. Reduction in MROs<br>51. No impact of COVID-19 on antibiotic resistance<br>52. No data available on impact of COVID-19 on antibiotic resistance<br>53. Suggested reasons for reduction in resistance<br>53.1. Reduced consumption<br>53.2. More hand washing/better IPC/distancing<br>53.3. Less screening due to lack of staff<br>53.4. Reduced reporting of antibiotic resistance |

| | |
|---|---|
| | 53.5. Fewer patients in hospital<br>53.6. Fewer samples available |
| Success stories | 54. Success stories during COVID-19 pandemic<br>54.1. Good responsiveness to AMR /COVID-19<br>54.2. Good coordination between actors to address COVID-19 and AMR<br>54.3. Improved IPC, hygiene, distancing<br>54.4. COVID-19 helped to identify gaps in IPC<br>54.5. Improved public understanding of infectious diseases and epidemiology<br>54.6. Antimicrobial stewardship integrated into COVID-19<br>54.7. Acquirement of equipment relevant to AMR<br>54.8. Delivery of training (on various topics)<br>54.9. Making something of AMR and COVID-19 to improve on both areas<br>54.10. Managed to continue activities/making improvements to AMR despite pandemic<br>54.11. Reduced antimicrobial consumption<br>54.12. Identification of areas needing to be improved in relation to AMR<br>54.13. COVID-19 has made it easier to control antibiotic prescribing<br>55. No success story available in relation to efforts to reduce AMR during COVID-19 pandemic |
| Prediction of long-term impacts | 56. Prediction of negative long-term impacts of COVID-19 on AMR<br>56.1. Increased prescribing/consumption of antibiotics<br>56.2. Increased use of non-prescribed antibiotics (self-prescribing)<br>56.3. Increased resistance of antibiotics /MDR<br>56.4. Inability to detect resistance (diagnostics<br>56.5. Less prioritisation of AMR projects/activities<br>56.6. Worsening of antimicrobial stewardship<br>56.7. Exposure to drug-resistant viruses<br>56.8. Less AMR surveillance<br>56.9. Staff fatigue and therefore unable to respond to AMR threats<br>56.10. Increased burden of infections<br>56.11. AMR threatens health systems<br>56.12. Financial tightening as result of COVID-19<br>56.13. Impact mainly on developing countries' economies and health<br>57. Prediction of positive long-term impacts of COVID-19 on AMR<br>57.1. Reduction in antibiotic consumption/improved rational use of antibiotics<br>57.2. Improved antimicrobial stewardship<br>57.3. Strengthening of One Health approach<br>57.4. Improved diagnostic capacity |

| | |
|---|---|
| | 57.5. Improved IPC<br>57.6. Improved global health system (preparedness)<br>57.7. Implementation of national action plans<br>57.8. Reduced/maintained resistance<br>57.9. Improved health-seeking behaviours<br>57.10. Improved surveillance<br>57.11. Improved awareness amongst decision makers of laboratories<br>57.12. Highlight AMR/epidemics as an issue<br>57.13. More focus on travel-associated infections in future<br>58. Prediction of long-term impacts of COVID-19 on AMR depends on duration of pandemic<br>59. No prediction of long-term impacts of COVID-19 on AMR |
| Suggestions for improvements | 60. Suggestions for improvements to the impact of COVID-19 on AMR activities<br>60.1. Activities to improve prevention of AMR<br>60.1.1. Focus on IPC across whole health system<br>60.1.2. Continue with biosafety and biosecurity<br>60.1.3. Ensure AMR is in all health policies<br>60.1.4. Increase resiliency of AMR programmes for future threats<br>60.1.5. Strengthen health systems<br>60.1.6. Better global coordinated resource mobilisation<br>60.1.7. Improve preparedness so pandemics have less impact on AMR<br>60.1.8. Improve compliance with AMR-relevant guidelines<br>60.1.9. Improve patient equity<br>60.2. Increased advocacy for AMR in order to receive more funding and resources<br>60.3. More public awareness of AMR and AMR-relevant areas<br>60.4. Focus on surveillance<br>60.5. Increased external support to countries<br>60.5.1. Support and engagement from WHO<br>60.5.1.1. On AMR & COVID-19<br>60.5.1.2. Capacity building for diagnostic facilities<br>60.5.2. Support with impacts of COVID-19 on AMR<br>60.5.3. Technical support<br>60.5.4. Financial support<br>60.5.5. Shift in external support from surveillance to research and development<br>60.5.6. Development of resources<br>60.5.6.1. Guidelines for the prudent use of antibiotics during a pandemic<br>60.5.6.2. National recommendations for treatment of suspected bacterial pneumonia at early stage<br>60.6. Increased resources for particular areas of AMR |

| | |
|---|---|
| | 60.6.1. IPC<br>60.6.2. Lab diagnostics<br>60.6.3. Surveillance<br>60.6.4. Antimicrobial stewardship<br>60.6.5. Laboratory supplies<br>60.6.6. Training of staff in particular AMR-relevant areas<br>60.6.7. Strengthened Water, Sanitation and Hygiene (WASH)<br>60.6.8. Research and development of new antibiotics<br>60.7. Focus on AMR in areas affected by COVID-19<br>60.7.1. Improved education about prescribing of antibiotics to patients with COVID-19<br>60.7.2. Training on IPC in COVID-19 settings<br>60.8. Support to improve quality of life of people who have had an infection<br>60.9. Technology and other advancements<br>60.9.1. Genomic surveillance of COVID-19 offers opportunities for AMR<br>60.9.2. Support more efficient ways of working (e.g. electronic prescribing and surveillance systems, use of artificial intelligence (AI))<br>60.10. Better communication<br>60.10.1. Between laboratory staff and physicians<br>60.10.2. More engagement between National Focal Point (NFP) and AMR teams in hospitals, including rewarding them for their work<br>60.11. Research/analysis<br>60.11.1. Survey on antimicrobial use in Sub-Saharan countries<br>60.11.2. Research on prescribing of antibiotics and antiviral for patients with COVID-19<br>60.11.3. Studies on risk factors and potential role of infections in COVID-19<br>60.11.4. Research opportunities for AMR<br>61. No suggestions for improvements to the impact of COVID-19 on AMR activities |
| Miscellaneous | 62. Country priorities<br>62.1. Find ways to make use of COVID-19 resources for AMR<br>62.2. Commitment to tackle AMR in their country<br>62.3. Training in staff awareness on AMR<br>62.4. Miscellaneous (includes regular auditing of antimicrobial use, monitoring of unnecessary use of antibiotics, better prescribing)<br>63. Miscellaneous impact of COVID-19 on AMR activities<br>63.1. The currently imposed sanctions in [country] are major threat to AMR work<br>64. AMR is having less of an impact now than at start |
